# Dementia and Frailty Impact Postoperative Care Trajectories and Burden among Older Adults Undergoing Radical Cystectomy for Bladder Cancer

**DOI:** 10.64898/2026.06.04.26354768

**Authors:** John Ernandez, Lingwei Xiang, Rachel Adler, John Hsu, Samir Shah, Dae Kim, Boris Gershman, Matthew Mossanen, Joel Weissman

## Abstract

**OBJECTIVE:** Bladder cancer (BC) is predominantly a disease of older, comorbid adults, and radical cystectomy (RC), which is the gold standard treatment, carries considerable morbidity. We sought to determine the impact of baseline dementia and frailty on the care trajectory beyond the immediate postoperative period. We hypothesized that frail patients and those with dementia undergoing RC for BC will have poorer care trajectories.

**METHODS AND MATERIALS:** We identified Medicare beneficiaries ≥ 66 years old who underwent RC for BC in 2017 with 12 months of pre- and post-RC enrollment. Frailty and dementia were characterized using validated, claims-based measures. Associations between baseline frailty and dementia with postoperative care trajectory outcomes were determined using Fine-Gray competing risk models.

**RESULTS:** We identified 3,600 beneficiaries of whom 11.6% were frail and 3.4% met criteria for dementia. Patients with dementia were more likely to be frail, comorbid, and not receive standard-of-care neoadjuvant chemotherapy. Frailty was independently associated with ≥ 2 transitions in care level after index discharge from RC and skilled nursing facility (SNF) admissions within 1 year of RC, exposure to intensive post-RC interventions, including dialysis and feeding tube placement, and poorer survival. Dementia remained associated with SNF admissions regardless of frailty level

**CONCLUSIONS:** Among a contemporary cohort of older adults undergoing RC for BC, preoperative dementia and frailty were independently associated with poorer care trajectory beyond the immediate postoperative period after RC. Our work highlights a role for preoperative geriatric assessment in identifying and optimizing patients at greatest risk.

## INTRODUCTION

The risk of postoperative morbidity increases with age and burden of baseline comorbidities.^1^ Persons living with dementia (PLWDs) are specifically at risk of in-hospital mortality, lengthier hospital stays, and non-home discharges after surgery.^2^ Furthermore, PLWDs are at higher risk of delirium^3^ and undergoing secondary interventions, such as feeding tube placement, despite such therapies having limited survival benefit among PLWDs.^4^ Similarly, patients with baseline frailty are at greater risk of mortality and morbidity after surgery compared to robust patients.^5–6^ Frailty is a syndrome of increased vulnerability to external stressors due to a reduced physiologic reserve.^7^ There is likely a complex, non-overlapping relationship between frailty and baseline comorbidities, including dementia.^8^

The impact of baseline cognitive impairment and frailty on postoperative outcomes remains especially relevant among older adults with bladder cancer (BC) undergoing radical cystectomy (RC). BC is overwhelmingly a disease of older adults, with a median age of diagnosis of 73 years and patients having an average of 8 comorbidities at time of diagnosis.^9^ RC is the gold-standard surgical treatment for muscle-invasive BC, and it is associated with high rates of morbidity.^10–11^

While frailty and cognitive impairment may impact immediate outcomes after RC,^6,12^ we do not understand how they may impact a patient’s trajectory of care outside of the perioperative period. Patients are largely willing to accept a high burden of inpatient care, though few wish to pursue care if it results in subsequent functional or cognitive impairment.^13^ Additionally, PLWDs are at risk of numerous transitions in care (TICs) after discharge (e.g., bidirectional transitions from community to inpatient hospitalization or facility), which contribute substantial burden to the patient and caregiver.^14^ This latter point is especially relevant for post-RC care, where older adults may struggle to manage a new diversion.

Herein, we seek to determine the impact of baseline frailty and dementia on the care trajectory among older adults undergoing RC for BC. We define the care trajectory to include multiple TICs, use of new mobility devices, and exposure to intensive interventions after discharge from index RC. We hypothesize that frailty and dementia will be associated with a greater number of transitions out of the community and exposure to invasive interventions after index RC, and those with both frailty and dementia will have the poorest outcomes.

## METHODS AND MATERIALS

### Cohort

We identified Medicare fee-for-service beneficiaries aged ≥66 years who underwent RC with a principal diagnosis of BC using the 2017 Medicare inpatient (IP) claims. Additionally, they were required to have 12-month continuous enrollment in Parts A and B prior to and after RC; those with continuous enrollment until death after RC were included. Beneficiaries with HMO enrollment were excluded.

### Covariates

Baseline demographic covariates at time of RC included age, race, gender, community-dwelling status, smoking status, comorbidity, rurality, and receipt of neoadjuvant chemotherapy (NAC). Date of birth (DOB), race, and gender were determined from the Master Beneficiary Summary File. Age was calculated from DOB and RC date in the IP claims. Community-dwelling status was determined using Minimum Data Set (MDS) file. Patients without an MDS assessment within 180 days prior to index admission were classified as community-dwelling. Smoking status was determined using relevant International Classification of Diseases (ICD)-10 codes with a 1-year lookback from index RC.^15^ Preoperative comorbidity was determined using the Elixhauser Comorbidity Index (ECI).^16^ Rurality was defined based on Rural-Urban Commuting Area (RUCA) codes. Neoadjuvant chemotherapy is the standard of care among patients with muscle-invasive BC.^17^ Receipt of NAC was determined using relevant Healthcare Common Procedure Coding System (HCPCS) codes. All available beneficiary data within one year prior to RC were used to determine ECI, smoking status, and receipt of NAC.

Patients with Alzheimer’s disease and related dementias (ADRD) before and at time of RC were identified using a validated algorithm for use with Medicare data with a 12-month lookback.^18^ Frailty was defined using the claims-based frailty index (CFI), which is a deficit accumulation frailty measure determined in the 12-month pre-RC period. The CFI has been validated using Medicare data for a variety of outcomes, including mortality, hospitalization, and discharge to skilled nursing facility (SNF). Frailty was defined as a CFI ≥0.25 based on prior literature.^19^

### Outcomes

We defined endpoints to capture the care trajectory that may occur after RC, including use of new mobility devices, multiple TICs, functional decline, SNF and nursing home (NH) admissions, and receipt of intensive interventions. Use of a new mobility device was defined as presence of a device in the 3 months after RC that was not present for 1 year preoperatively. Mobility devices included use of cane, walker, wheelchair, or scooter and were determined using HCPCS files. Patients who died within 90 days of RC and without a new mobility device were excluded from this outcome. Multiple TICs were defined as ≥2 transitions in care level from the baseline level on discharge from RC for the subsequent year.

Patients who died after a single TIC were considered to meet criteria, though those who died within 1 year without any TIC were excluded. Admission and discharge data for each TIC were determined using SNF, MDS, Home Health Agency (HHA), and IP claims. Functional decline was defined as a change in CFI 12 months after index RC versus preoperative CFI. SNF and NH admissions among community-dwelling patients and NH-dwelling patients were determined at 30, 90, 180, and 365 days after index RC via IP, SNF, MDS, and HHA files. Patients who died prior to SNF/NH admission were excluded from this outcome. Intensive interventions within 90 days of RC were determined and included cardiopulmonary resuscitation, mechanical ventilation, intubation, prolonged renal replacement therapy, extracorporeal membrane oxygenation, and placement of a feeding tube.^20^ Any negative event within 365 days of index RC served as a composite measure of all above outcomes. Length of stay (LOS) was also recorded as days from RC to discharge. Survival was also determined at 30, 90, 180, and 365 days from index RC. Refer to Supplementary documents for relevant ICD, CPT, and HCPCS codes.

### Statistical Analysis

Baseline demographic covariates were summarized using medians and interquartile ranges (IQR) or frequency counts with percentages. Outcomes were stratified by ADRD status and frailty and compared across groups using the Wilcoxon two-sample test, Kruskal-Wallis test, Fisher’s Exact test, or chi-square test, as appropriate. Associations between outcomes and baseline covariates were evaluated using univariable and multivariable generalized estimating logistic equations with results summarized with odds ratios (ORs) with 95% confidence intervals (95% CIs). Outcomes modelled as either dichotomous or continuous, in the case of LOS. ADRD and frailty were also treated as four-level variables with ADRD/non-frail, non-ADRD/frail, and ADRD/frail cohorts each compared against a non-ADRD/non-frail cohort for each regression model. Multivariable regression was adjusted for baseline covariates, including age, race, gender, smoking status, ECI, rurality, and receipt of NAC. Survival analysis was conducted using Cox Proportional Hazard models, and all other outcomes were modelled via Fine-Gray method for consideration of death as a competing event with results summarized with hazard ratios (HRs) with 95% CIs. Hospital clustering was accounted in all models. Statistical analyses were performed using SAS, version 9.4 (SAS Institute Inc). All tests were two-sided with *p*-values <0.05 considered statistically significant.

## RESULTS

### Baseline Characteristics

A total of 3,600 beneficiaries were included in our analysis with baseline demographics summarized in Table 1. Median age was 74.9 (70.7 – 79.6) with 3,496 (97.1%) beneficiaries being community-dwelling at time of RC, 2,437 (67.7%) being current or former smokers, and 976 (27.1%) having receipt of NAC. Median ECI was 14 (7 – 31), and median CFI was 0.18 (0.15 – 0.21) with 419 (11.6%) beneficiaries meeting criteria for preoperative frailty. 122 beneficiaries (3.4%) met criteria for ADRD. Compared to those without ADRD, those with ADRD were significantly more likely to be older (78.8 versus 74.8 years; *p* < 0.0001), NH-dwelling (13.1 versus 2.5%; *p* < 0.0001), frail (67.2 versus 9.7%; *p* < 0.0001), current or former smokers (76.2 versus 67.4%; *p* = 0.0403), and comorbid (22.5 versus 14; *p* = 0.0476). Frail beneficiaries were also significantly more likely to have baseline ADRD (19.6 versus 1.3%, *p* < 0.0001). Those with ADRD status were less likely to have received NAC (14.8 versus 27.5%; *p* = 0.0018). There were no statistically significant differences in race, gender, or rurality based on ADRD status.

**Table 1.**
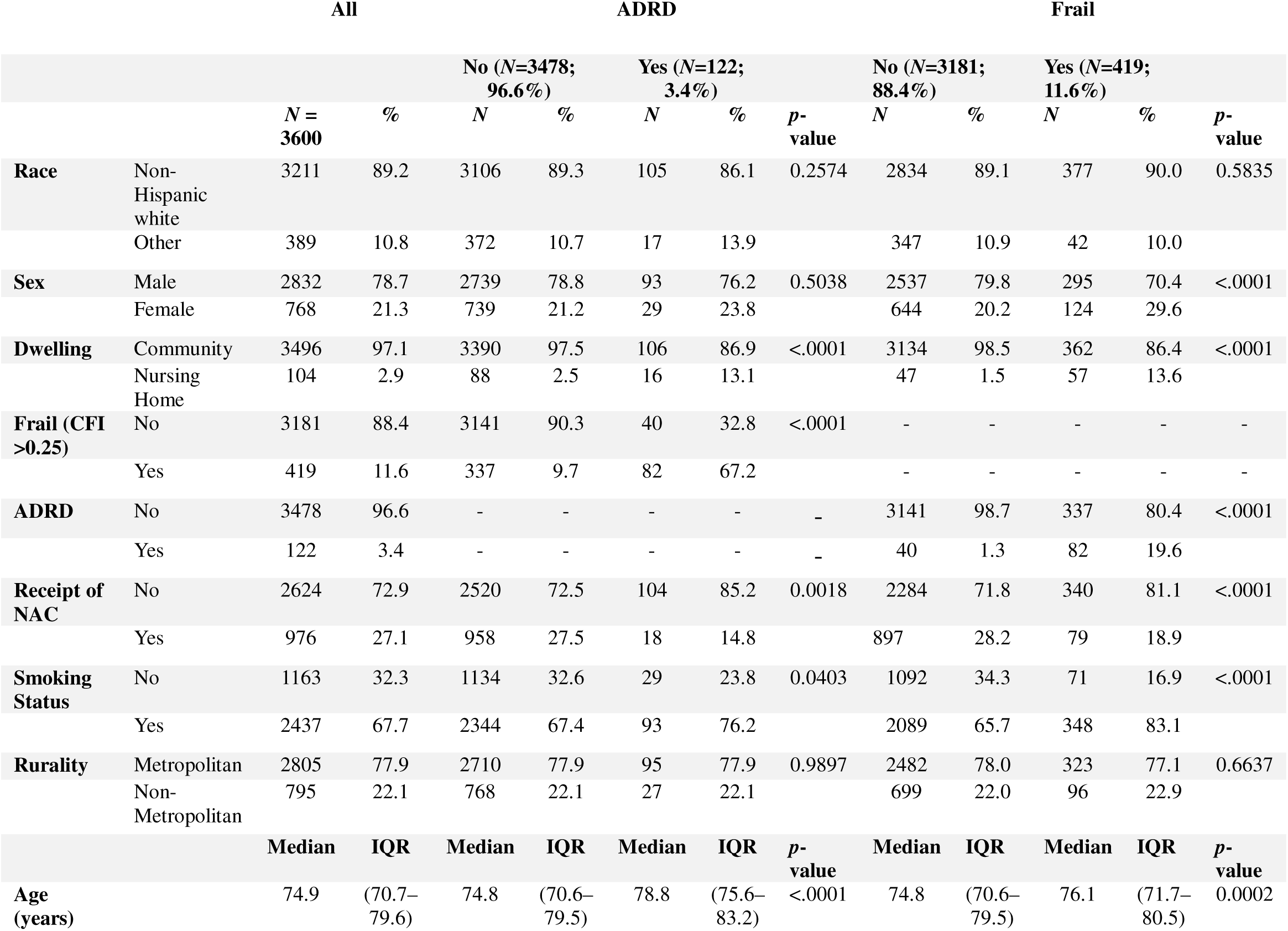
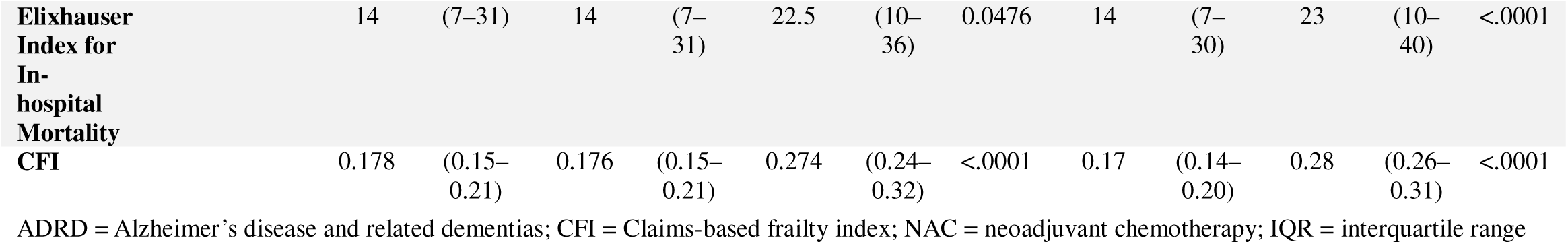
Baseline sociodemographic covariates stratified by ADRD status.

|  |  | All |  | ADRD |  |  |  | Frail |  |  |  |  |  |
| --- | --- | --- | --- | --- | --- | --- | --- | --- | --- | --- | --- | --- | --- |
|  |  |  |  | No (N=3478;<br>96.6%) |  | Yes (N=122;<br>3.4%) |  | p-value | No (N=3181;<br>88.4%) |  | Yes (N=419;<br>11.6%) |  | p-value |
|  |  | N =<br>3600 | % | N | % | N | % |  | N | % | N | % |  |
| Race | Non-Hispanic white | 3211 | 89.2 | 3106 | 89.3 | 105 | 86.1 | 0.2574 | 2834 | 89.1 | 377 | 90.0 | 0.5835 |
|  | Other | 389 | 10.8 | 372 | 10.7 | 17 | 13.9 |  | 347 | 10.9 | 42 | 10.0 |  |
| Sex | Male | 2832 | 78.7 | 2739 | 78.8 | 93 | 76.2 | 0.5038 | 2537 | 79.8 | 295 | 70.4 | <.0001 |
|  | Female | 768 | 21.3 | 739 | 21.2 | 29 | 23.8 |  | 644 | 20.2 | 124 | 29.6 |  |
| Dwelling | Community | 3496 | 97.1 | 3390 | 97.5 | 106 | 86.9 | <.0001 | 3134 | 98.5 | 362 | 86.4 | <.0001 |
|  | Nursing Home | 104 | 2.9 | 88 | 2.5 | 16 | 13.1 |  | 47 | 1.5 | 57 | 13.6 |  |
| Frail (CFI >0.25) | No | 3181 | 88.4 | 3141 | 90.3 | 40 | 32.8 | <.0001 | - | - | - | - | - |
|  | Yes | 419 | 11.6 | 337 | 9.7 | 82 | 67.2 |  | - | - | - | - | - |
| ADRD | No | 3478 | 96.6 | - | - | - | - | - | 3141 | 98.7 | 337 | 80.4 | <.0001 |
|  | Yes | 122 | 3.4 | - | - | - | - | - | 40 | 1.3 | 82 | 19.6 |  |
| Receipt of NAC | No | 2624 | 72.9 | 2520 | 72.5 | 104 | 85.2 | 0.0018 | 2284 | 71.8 | 340 | 81.1 | <.0001 |
|  | Yes | 976 | 27.1 | 958 | 27.5 | 18 | 14.8 |  | 897 | 28.2 | 79 | 18.9 |  |
| Smoking Status | No | 1163 | 32.3 | 1134 | 32.6 | 29 | 23.8 | 0.0403 | 1092 | 34.3 | 71 | 16.9 | <.0001 |
|  | Yes | 2437 | 67.7 | 2344 | 67.4 | 93 | 76.2 |  | 2089 | 65.7 | 348 | 83.1 |  |
| Rurality | Metropolitan | 2805 | 77.9 | 2710 | 77.9 | 95 | 77.9 | 0.9897 | 2482 | 78.0 | 323 | 77.1 | 0.6637 |
|  | Non-Metropolitan | 795 | 22.1 | 768 | 22.1 | 27 | 22.1 |  | 699 | 22.0 | 96 | 22.9 |  |
|  |  | Median | IQR | Median | IQR | Median | IQR | p-value | Median | IQR | Median | IQR | p-value |
| Age (years) |  | 74.9 | (70.7–79.6) | 74.8 | (70.6–79.5) | 78.8 | (75.6–83.2) | <.0001 | 74.8 | (70.6–79.5) | 76.1 | (71.7–80.5) | 0.0002 |

|  |  |  |  |  |  |  |  |  |  |  |  |  |
| --- | --- | --- | --- | --- | --- | --- | --- | --- | --- | --- | --- | --- |
| **Elixhauser**<br>**Index for**<br>**In-**<br>**hospital**<br>**Mortality** | 14 | (7–31) | 14 | (7–31) | 22.5 | (10–36) | 0.0476 | 14 | (7–30) | 23 | (10–40) | <.0001 |
| **CFI** | 0.178 | (0.15–0.21) | 0.176 | (0.15–0.21) | 0.274 | (0.24–0.32) | <.0001 | 0.17 | (0.14–0.20) | 0.28 | (0.26–0.31) | <.0001 |

ADRD = Alzheimer's disease and related dementias; CFI = Claims-based frailty index; NAC = neoadjuvant chemotherapy; IQR = interquartile range

Frequency of events are summarized in Table 2. Among beneficiaries with ADRD, there were significant differences in the frequency of several outcomes, including: multiple TICs (occurring in 60.9% of those with ADRD versus 44.1% without ADRD; *p* = 0.0019); SNF admission up to 365 days (61.4 vs 29.1%, *p* < 0.0001); NH admission at 365 days (16.4 vs 4.9%, *p* = 0.0005); and mortality up to 365 days (48.4 vs. 23.9%; *p* < 0.0001). Among frail beneficiaries, there were significant differences in the frequency of several outcomes, including: multiple TICs (66.0 vs. 42.2%, *p* < 0.0001); experiencing any intensive intervention within 90 days of RC (11.8 vs. 3.5%; *p* < 0.0001); SNF admission up to 365 days (58.8 vs. 26.9%, *p* < 0.0001); NH admissions up to 365 days (11.4 vs. 4.6%, *p* < 0.0001); mortality up to 365 days (44.2 vs. 22.1%, *p* < 0.0001); and any negative event within 365 days of RC (88.5 vs 81.4%, *p* = 0.0003). Frail beneficiaries with ADRD generally were smaller cohorts, though had significantly higher incidence of TIC, SNF admission, and mortality than robust/non-ADRD beneficiaries. Beneficiaries with baseline frailty (47.4 versus 72.3%, *p* < 0.0001) or ADRD (54.3 versus 70.0%, *p* = 0.0003) were less likely to experience functional decline. There were no significant differences in mobility device use.

**Table 2.** Frequency of study outcomes stratified by ADRD and frailty status.

|  | All |  | ADRD |  |  |  |  | Frail |  |  |  |  | Both |  |  |  |  |
| --- | --- | --- | --- | --- | --- | --- | --- | --- | --- | --- | --- | --- | --- | --- | --- | --- | --- |
|  |  |  | No |  | Yes |  | p-value | No |  | Yes |  | p-value | ADRD & Frail |  | No ADRD/Robust |  | p-value |
| Outcome | N | % | N | % | N | % |  | N | % | N | % |  | N | % | N | % |  |
| New mobility device (N=3309) | 516 | 15.6 | 499 | 15.5 | 17 | 18.9 | 0.3823 | 455 | 15.3 | 61 | 18.4 | 0.134 | * | * | 450 | 15.3 | 0.4376 |
| TIC (N=3220) | 1436 | 44.6 | 1383 | 44.1 | 53 | 60.9 | 0.0019 | 1224 | 42.2 | 212 | 66.0 | <.0001 | 38 | 71.7 | 1209 | 42.2 | <.0001 |
| Functional decline (N=3527) | 2451 | 69.5 | 2388 | 70.0 | 63 | 54.3 | 0.0003 | 2263 | 72.3 | 188 | 47.4 | <.0001 | 38 | 50 | 2238 | 72.4 | <.0001 |
| 30-day SNF admission, among community-dwelling patients (N=3376) | 707 | 20.9 | 661 | 20.2 | 46 | 46.5 | <.0001 | 567 | 18.6 | 140 | 42.4 | <.0001 | 30 | 50.9 | 551 | 18.3 | <.0001 |
| 90-day SNF admission (N=3286) | 795 | 24.2 | 746 | 23.4 | 49 | 52.1 | <.0001 | 637 | 21.4 | 158 | 51.5 | <.0001 | 32 | 58.2 | 620 | 21.1 | <.0001 |
| 180-day SNF admission (N=3177) | 834 | 26.3 | 785 | 25.5 | 49 | 52.7 | <.0001 | 672 | 23.4 | 162 | 54 | <.0001 | 32 | 59.3 | 655 | 23.1 | <.0001 |
| 365-day SNF admission (N=2985) | 896 | 30.0 | 842 | 29.1 | 54 | 61.4 | <.0001 | 726 | 26.9 | 170 | 58.8 | <.0001 | 36 | 69.2 | 708 | 26.6 | <.0001 |
| 30-day NH admission, among community-dwelling patients (N=3347) | 42 | 1.3 | * | * | * | * | 0.0067 | 31 | 1.0 | 11 | 3.4 | 0.0016 | * | * | 29 | 1.0 | 0.0001 |
| 90-day NH admission (N=3177) | 78 | 2.5 | * | * | * | * | 0.0008 | 60 | 2.1 | 18 | 6.5 | <.0001 | * | * | 57 | 2.0 | <.0001 |
| 180-day NH admission (N=2996) | 108 | 3.6 | * | * | * | * | 0.0058 | 87 | 3.2 | 21 | 8.1 | <.0001 | * | * | 84 | 3.1 | <.0001 |
| 365-day NH admission (N=2726) | 141 | 5.2 | 130 | 4.9 | 11 | 16.4 | 0.0005 | 115 | 4.6 | 26 | 11.4 | <.0001 | * | * | 111 | 4.5 | <.0001 |
| 30-day SNF admission, among NH patients (N=91) | 57 | 62.6 | * | * | * | * | 0.7571 | 25 | 61 | 32 | 64 | 0.7667 | * | * | 25 | 61.0 | 0.8587 |
| 90-day SNF admission (N=86) | 60 | 69.8 | * | * | * | * | 0.7169 | 27 | 65.9 | 33 | 73.3 | 0.4506 | * | * | 27 | 65.9 | 0.7138 |
| 180-day SNF admission (N=83) | 61 | 73.5 | * | * | * | * | - | * | * | * | * | 0.2888 | * | * | 28 | 68.3 | 0.5687 |
| 365-day SNF admission (N=79) | 64 | 81.0 | * | * | * | * | - | * | * | * | * | 0.0649 | * | * | 30 | 73.2 | 0.1797 |
| 30-day mortality (N=3600) | 124 | 3.4 | 112 | 3.2 | 12 | 9.8 | <.0001 | 87 | 2.7 | 37 | 8.8 | <.0001 | * | * | 85 | 2.7 | <.0001 |
| 90-day mortality (N=3600) | 323 | 9.0 | 289 | 8.3 | 34 | 27.9 | <.0001 | 229 | 7.2 | 94 | 22.4 | <.0001 | * | * | 222 | 7.1 | <.0001 |
| 180-day mortality (N=3600) | 563 | 15.6 | 519 | 14.9 | 44 | 36.1 | <.0001 | 421 | 13.2 | 142 | 33.9 | <.0001 | * | * | 413 | 13.2 | <.0001 |
| 365-day mortality (N=3600) | 889 | 24.7 | 830 | 23.9 | 59 | 48.4 | <.0001 | 704 | 22.1 | 185 | 44.2 | <.0001 | 47 | 57.3 | 692 | 22.0 | <.0001 |
| 90-day intensive intervention (N=3314) | 143 | 4.3 | * | * | * | * | 0.0165 | 103 | 3.5 | 40 | 11.8 | <.0001 | * | * | 101 | 3.4 | <.0001 |
| Any negative event (N=3600) | 2961 | 82.3 | 2854 | 82.1 | 107 | 87.7 | 0.1086 | 2590 | 81.4 | 371 | 88.5 | 0.0003 | * | * | 2559 | 81.5 | 0.0023 |
|  | **Median** | **IQR** | **Median** | **IQR** | **Median** | **IQR** | **p-value** | **Median** | **IQR** | **Median** | **IQR** | **p-value** | **Median** | **IQR** | **Median** | **IQR** | **p-value** |
| LOS | 7 | (5,9) | 7 | (5,9) | 7 | (5,11) | 0.2414 | 7 | (5,9) | 8 | (6,14) | <.0001 | 7 | (6,13) | 7 | (5,9) | <.0001 |
| Change in CFI (after RC – before RC) | 0.025 | (-0.007, 0.064) | 0.025 | (-0.007, 0.064) | 0.008 | (-0.031, 0.062) | 0.0025 | 0.028 | (-0.004, 0.066) | -0.004 | (-0.048, 0.041) | <0.001 | -0.001 | (-0.056, 0.044) | 0.028 | (-0.004, 0.066) | <0.001 |

ADRD = Alzheimer's disease and related dementias; TIC = transitions in care; SNF = skilled nursing facility; NH = nursing home; IQR = interquartile range. \* = Censored per CMS cell suppression policy.

### Unadjusted Outcomes

Univariable associations of ADRD status and frailty with each outcome are summarized in Supplementary Table 2a-b. Beneficiaries who met criteria for ADRD were more likely have multiple TICs (OR 2.00; 95% CI 1.25 – 3.11; *p* = 0.0034); SNF admission up to 365 days after RC (OR 3.88; 95% CI 2.51 – 6.00; *p* < 0.0001); NH admission up to 365 days after RC (OR 3.82; 95% CI 1.80 – 8.13; *p* = 0.0005); experience any intensive intervention within 90 days of RC (OR 2.50; 95% CI 1.27 – 4.92; *p* = 0.0081); and mortality after RC up to 365 days (OR 2.99; 95% CI 2.10 – 4.26; *p* < 0.0001). Frail, community-dwelling beneficiaries were more likely to have multiple TICs (OR 2.66; 95% CI 2.08 – 3.41; *p* < 0.0001); SNF admission up to 365 days after RC (OR 3.88; 95% CI 2.98 – 5.05; *p* < 0.0001); NH admission up to 365 days after RC (OR 2.66; 95% CI 1.62 – 4.38; *p* = 0.0001); experience intensive interventions at 90 days (OR 3.74; 95% CI 2.56 – 5.48; *p* < 0.0001); greater LOS (OR 3.23; 95% CI 2.25 – 4.21; *p* < 0.0001); mortality up to 365 days after RC (OR 2.78; 95% CI 2.30 – 3.37; *p* < 0.0001); and experience any negative event within 365 days from RC (OR 1.76; 95% CI 1.31 – 2.37; *p* = 0.0002).

### Adjusted Outcomes

Multivariable associations of ADRD status and frailty with each outcome are summarized in Table 3a-b. Beneficiaries who met criteria for ADRD were more likely to have multiple TICs (OR 1.73; 95% CI 1.09 – 2.75; p = 0.0207); SNF admission up to 365 days after RC (OR 2.79; 95% CI 2.79 – 4.42; *p* < 0.0001); NH admission up to 365 days (OR 3.12; 95% CI 1.42 – 6.84; *p* = 0.0045); and mortality up to 365 days after RC (OR 2.37; 95% CI 1.64 – 3.43; *p* < 0.0001). After adjustment, ADRD status was not associated with 90-day intensive interventions. Frail beneficiaries were more likely to have multiple TICs (OR 2.37; 95% CI 1.85 – 3.04; *p* < 0.0001), SNF admission up to 365 days after RC (OR 3.31; 95% CI 2.45 – 4.47; *p* < 0.0001); NH admission up to 365 days after RC (OR 2.37; 95% CI 1.40 – 4.03; *p* = 0.0014); experience intensive interventions within 90 days of RC (OR 2.81; 95% CI 1.82 – 4.32; *p* < 0.0001); greater LOS (OR 2.75; 95% CI 1.83 – 3.70; *p* < 0.0001); and mortality up to 365 days after RC (OR 2.11; 95% CI 1.70 – 2.62; *p* < 0.0001). Adjusted associations with multiple TICs, SNF and NH admissions, exposure to intensive interventions, and survival were maintained among ADRD/frail beneficiaries when modeling ADRD and frailty as a four-level variable (see Table 3b). Similarly, there remained significant associations after adjustment between frailty and multiple TICs, SNF and NH admission, exposure to intensive interventions, and overall survival after accounting for competing risk of death. A similar association remained between ADRD and SNF admission and overall survival on adjusted competing risk analysis (see Supplementary Table 4a).

**Table 3a.** Multivariable adjusted associations between ADRD status or frailty and outcomes.

| Outcome | ADRD |  |  | Frail |  |  |
| --- | --- | --- | --- | --- | --- | --- |
|  | OR | 95% CI | p-value | OR | 95% CI | p-value |
| New mobility device (N=3309) | 1.12 | 0.66, 1.90 | 0.684 | 1.13 | 0.82, 1.55 | 0.447 |
| TIC (N=3220) | 1.73 | 1.09, 2.75 | 0.021 | 2.37 | 1.85, 3.04 | <.0001 |
| Functional Decline (N=3527) | 0.49 | 0.33, 0.73 | 0.000 | 0.34 | 0.28, 0.42 | <.0001 |
| 30-day SNF admission, among community-dwelling patients (N=3376) | 2.44 | 1.56, 3.83 | <.0001 | 2.65 | 1.98, 3.55 | <.0001 |
| 90-day SNF admission (N=3286) | 2.51 | 2.51, 4.00 | 0.000 | 3.29 | 2.45, 4.43 | <.0001 |
| 180-day SNF admission (N=3177) | 2.26 | 1.42, 3.61 | 0.001 | 3.24 | 2.42, 4.36 | <.0001 |
| 365-day SNF admission (N=2985) | 2.79 | 2.80, 4.42 | <.0001 | 3.31 | 2.46, 4.47 | <.0001 |
| 30-day NH admission, among community-dwelling patients (N=3347) | 3.53 | 1.05, 11.92 | 0.042 | 3.20 | 1.32, 7.76 | 0.010 |
| 90-day NH admission (N=3177) | 3.42 | 1.30, 9.03 | 0.013 | 2.79 | 1.38, 5.63 | 0.004 |
| 180-day NH admission (N=2996) | 2.50 | 0.99, 6.31 | 0.052 | 2.34 | 1.29, 4.24 | 0.005 |
| 365-day NH admission (N=2726) | 3.12 | 1.42, 6.84 | 0.005 | 2.37 | 1.40, 4.03 | 0.001 |
| 30-day SNF admission, among NH patients (N=91) | 0.27 | 0.06, 1.12 | 0.071 | 1.30 | 0.51, 3.27 | 0.583 |
| 90-day SNF admission (N=86) | 0.45 | 0.07, 3.11 | 0.421 | 1.35 | 0.50, 3.65 | 0.549 |
| 180-day SNF admission (N=83) | 0.41 | 0.06, 2.93 | 0.376 | 1.97 | 0.67, 5.83 | 0.222 |
| 365-day SNF admission (N=79) | 0.29 | 0.02, 3.81 | 0.346 | 3.95 | 0.93, 16.75 | 0.063 |
| 30-day mortality (N=3600) | 2.21 | 1.14, 4.27 | 0.019 | 2.63 | 1.66, 4.17 | <.0001 |
| 90-day mortality (N=3600) | 3.16 | 2.0, 5.01 | <.0001 | 2.65 | 1.98, 3.55 | <.0001 |
| 180-day mortality (N=3600) | 2.48 | 1.65, 3.72 | <.0001 | 2.49 | 1.97, 3.15 | <.0001 |
| 365-day mortality (N=3600) | 2.37 | 1.64, 3.43 | <.0001 | 2.11 | 1.70, 2.62 | <.0001 |
| 90-day intensive intervention (N=3314) | 1.93 | 0.95, 3.91 | 0.070 | 2.81 | 1.82, 4.32 | <.0001 |
| Any negative event (N=3600) | 1.25 | 0.72, 2.17 | 0.434 | 1.49 | 1.09, 2.02 | 0.011 |
| LOS* | 0.50 | -0.82, 1.82 | 0.455 | 2.75 | 1.83, 3.66 | <.0001 |

ADRD = Alzheimer's disease and related dementias; TIC = transitions in care; SNF = skilled nursing facility; NH = nursing home; LOS = length of stay. Reference groups are non-ADRD and non-frail beneficiaries. \*Outcomes for LOS reflects mean difference, not OR.

**Table 3b.**
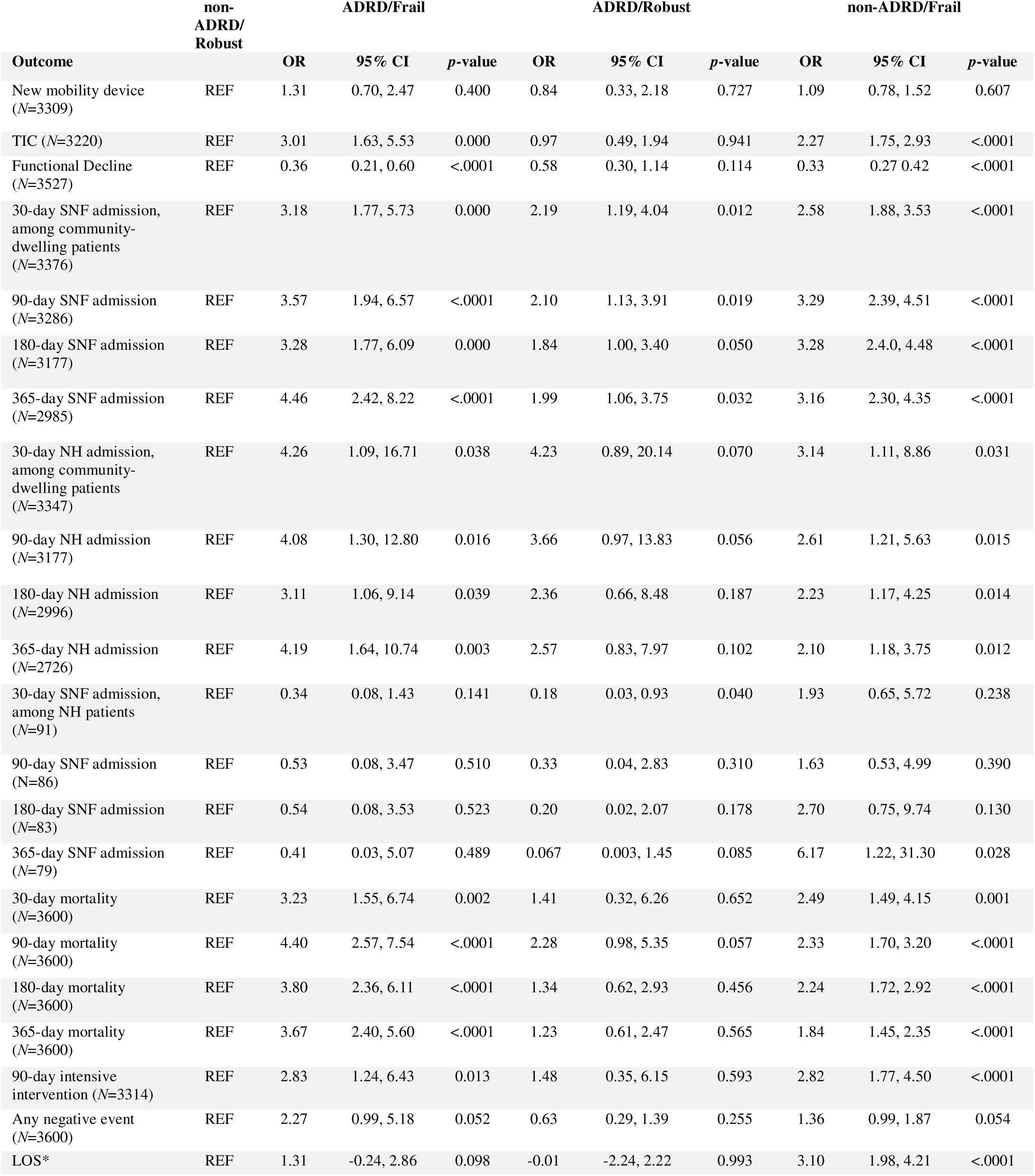
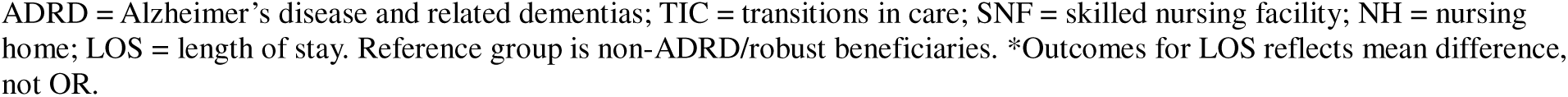
Multivariable adjusted associations between four-level ADRD/frailty status and outcomes.

| Outcome | non-ADRD/<br>Robust | ADRD/Frail |  |  | ADRD/Robust |  |  | non-ADRD/Frail |  |  |
| --- | --- | --- | --- | --- | --- | --- | --- | --- | --- | --- |
|  |  | OR | 95% CI | p-value | OR | 95% CI | p-value | OR | 95% CI | p-value |
| New mobility device (N=3309) | REF | 1.31 | 0.70, 2.47 | 0.400 | 0.84 | 0.33, 2.18 | 0.727 | 1.09 | 0.78, 1.52 | 0.607 |
| TIC (N=3220) | REF | 3.01 | 1.63, 5.53 | 0.000 | 0.97 | 0.49, 1.94 | 0.941 | 2.27 | 1.75, 2.93 | <.0001 |
| Functional Decline (N=3527) | REF | 0.36 | 0.21, 0.60 | <.0001 | 0.58 | 0.30, 1.14 | 0.114 | 0.33 | 0.27 0.42 | <.0001 |
| 30-day SNF admission, among community-dwelling patients (N=3376) | REF | 3.18 | 1.77, 5.73 | 0.000 | 2.19 | 1.19, 4.04 | 0.012 | 2.58 | 1.88, 3.53 | <.0001 |
| 90-day SNF admission (N=3286) | REF | 3.57 | 1.94, 6.57 | <.0001 | 2.10 | 1.13, 3.91 | 0.019 | 3.29 | 2.39, 4.51 | <.0001 |
| 180-day SNF admission (N=3177) | REF | 3.28 | 1.77, 6.09 | 0.000 | 1.84 | 1.00, 3.40 | 0.050 | 3.28 | 2.4.0, 4.48 | <.0001 |
| 365-day SNF admission (N=2985) | REF | 4.46 | 2.42, 8.22 | <.0001 | 1.99 | 1.06, 3.75 | 0.032 | 3.16 | 2.30, 4.35 | <.0001 |
| 30-day NH admission, among community-dwelling patients (N=3347) | REF | 4.26 | 1.09, 16.71 | 0.038 | 4.23 | 0.89, 20.14 | 0.070 | 3.14 | 1.11, 8.86 | 0.031 |
| 90-day NH admission (N=3177) | REF | 4.08 | 1.30, 12.80 | 0.016 | 3.66 | 0.97, 13.83 | 0.056 | 2.61 | 1.21, 5.63 | 0.015 |
| 180-day NH admission (N=2996) | REF | 3.11 | 1.06, 9.14 | 0.039 | 2.36 | 0.66, 8.48 | 0.187 | 2.23 | 1.17, 4.25 | 0.014 |
| 365-day NH admission (N=2726) | REF | 4.19 | 1.64, 10.74 | 0.003 | 2.57 | 0.83, 7.97 | 0.102 | 2.10 | 1.18, 3.75 | 0.012 |
| 30-day SNF admission, among NH patients (N=91) | REF | 0.34 | 0.08, 1.43 | 0.141 | 0.18 | 0.03, 0.93 | 0.040 | 1.93 | 0.65, 5.72 | 0.238 |
| 90-day SNF admission (N=86) | REF | 0.53 | 0.08, 3.47 | 0.510 | 0.33 | 0.04, 2.83 | 0.310 | 1.63 | 0.53, 4.99 | 0.390 |
| 180-day SNF admission (N=83) | REF | 0.54 | 0.08, 3.53 | 0.523 | 0.20 | 0.02, 2.07 | 0.178 | 2.70 | 0.75, 9.74 | 0.130 |
| 365-day SNF admission (N=79) | REF | 0.41 | 0.03, 5.07 | 0.489 | 0.067 | 0.003, 1.45 | 0.085 | 6.17 | 1.22, 31.30 | 0.028 |
| 30-day mortality (N=3600) | REF | 3.23 | 1.55, 6.74 | 0.002 | 1.41 | 0.32, 6.26 | 0.652 | 2.49 | 1.49, 4.15 | 0.001 |
| 90-day mortality (N=3600) | REF | 4.40 | 2.57, 7.54 | <.0001 | 2.28 | 0.98, 5.35 | 0.057 | 2.33 | 1.70, 3.20 | <.0001 |
| 180-day mortality (N=3600) | REF | 3.80 | 2.36, 6.11 | <.0001 | 1.34 | 0.62, 2.93 | 0.456 | 2.24 | 1.72, 2.92 | <.0001 |
| 365-day mortality (N=3600) | REF | 3.67 | 2.40, 5.60 | <.0001 | 1.23 | 0.61, 2.47 | 0.565 | 1.84 | 1.45, 2.35 | <.0001 |
| 90-day intensive intervention (N=3314) | REF | 2.83 | 1.24, 6.43 | 0.013 | 1.48 | 0.35, 6.15 | 0.593 | 2.82 | 1.77, 4.50 | <.0001 |
| Any negative event (N=3600) | REF | 2.27 | 0.99, 5.18 | 0.052 | 0.63 | 0.29, 1.39 | 0.255 | 1.36 | 0.99, 1.87 | 0.054 |
| LOS* | REF | 1.31 | -0.24, 2.86 | 0.098 | -0.01 | -2.24, 2.22 | 0.993 | 3.10 | 1.98, 4.21 | <.0001 |

ADRD = Alzheimer's disease and related dementias; TIC = transitions in care; SNF = skilled nursing facility; NH = nursing home; LOS = length of stay. Reference group is non-ADRD/robust beneficiaries. \*Outcomes for LOS reflects mean difference, not OR.

## DISCUSSION

Bladder cancer is largely a disease of older, comorbid adults.^9^ Radical cystectomy, which is the gold standard treatment for muscle-invasive bladder cancer, carries considerable morbidity.^10–11^ Frailty and dementia are both associated with poorer outcomes after cystectomy,^6,12^ though it remains unclear how these conditions impact care trajectories beyond the immediate postoperative period. We found patients with dementia undergoing RC were at greater risk of SNF and NH admission, multiple transitions in care levels, and poorer survival up to one year from RC. Frail older adults undergoing RC were additionally at higher risk of intensive postoperative interventions, suggesting that the effects of frailty and dementia extend beyond the perioperative period. While cohorts were small, those with dementia who were also frail were similarly at greater risk of poorer outcomes, though it appears their outcomes were largely driven by frailty. Our study has several strengths; namely, we report the first study to our knowledge to describe the impact of preoperative dementia and frailty, as well as their interactions, on older adults undergoing RC for BC. We utilize a contemporary, national cohort of older adults, validated measures of both dementia and frailty, and novel outcomes designed to capture burden of care beyond the immediate perioperative period.

We found that 3.4% of our cohort of older adults met preoperative criteria for ADRD. The incidence of dementia among older adults undergoing RC has not been well established. Jazzar, *et al.* reported 17.4% of patients developing ADRD after treatment for BC, though their cohort includes adults undergoing non-operative management of BC and does not capture preoperative incidence.^21^ Grunewald, *et al.* prospectively screened patients undergoing RC and found 27% (14/51) met criteria for mild cognitive impairment, though this is not necessarily comparable to more severe dementia.^12^ Patients with ADRD in our study were more likely to be older, frailer, and more comorbid, which has been demonstrated in prior surgical cohorts.^1–2^ Our study found that those with ADRD were also less likely to receive neoadjuvant chemotherapy, which is standard-of-care practice in treatment of muscle-invasive BC.^17^ Given that those with ADRD in our cohort were significantly more comorbid, they may have been deemed unfit for chemotherapy. Additionally, 11.6% of older adults in our cohort were mildly frail or greater. The reported prevalence of frailty among patients undergoing RC for BC varies considerably with frailty measures utilized, though has been reported as high as 48.9% for patients >70 years old.^6,22^ Gershman, *et al.* reported 7% of older adults in a Medicare cohort undergoing RC were at least mildly frail as defined by CFI.^23^

It is well-established that preoperative dementia is associated with risk of non-home discharge after elective and emergent surgery.^3^ However, our work is the first to our knowledge to demonstrate that older adults with BC and ADRD undergoing RC are at increased risk of SNF and NH admission, independent of comorbidity and frailty; this was observed in 61.4% and 16.4% of older adults with ADRD in our cohort by 1 year from RC, respectively. These findings may be due to the impact of preoperative dementia on postoperative delirium and subsequent cognitive decline. Preoperative dementia has been shown to increase risk of postoperative delirium.^4^ Large, *et al.* demonstrated that those with BC undergoing RC who developed delirium were also more likely to utilize postoperative care.^24^ Preoperative dementia has also been shown to increase risk of postoperative cognitive decline.^25^ After RC, patients may be required to manage a urostomy and surgical drains, self-catheterize a continent diversion, and participate in rehabilitation. Accordingly, caregivers of patients with dementia may advocate for non-home discharge after RC given concerns with engaging in care. Our study demonstrates that those with ADRD continue to be at increased risk of SNF admission one year from RC, which may highlight the effects of ongoing cognitive decline.

We found that frail adults undergoing RC were at increased risk of two or more transitions in care level within a year of RC, including SNF and NH admission. We suspect transitions in care are driven by complication-related care utilization amplified by poor physiologic reserve among frail patients. Urinary diversion with small bowel carries considerable risk of metabolic acidosis and urosepsis.^26^ Frailer patients undergoing RC are at greater risk of these complications, with 73% undergoing a complication requiring care utilization within 90 days of RC.^23^ Multiple transitions in care level also create possibilities for medical error and generate financial burden for patients and caregivers, let alone baseline treatment-related burden which frail adults undergoing BC treatment are more likely to report.^14,28^ Similarly, those with ADRD were at greater risk of multiple transitions in care after RC, occurring in 60.9% of those with ADRD in our cohort within one year of RC. It remains imperative to determine the utility of a SNF or NH admission for PLWDs transitioning from the community after RC beyond addressing their postoperative care needs given the risk of subsequent institutionalization.^14^

Frail adults were also at risk of undergoing intensive interventions within 90 days of RC, which was observed in 11.8% of frail patients and 4.3% of the overall cohort. Incidence of intensive interventions after RC are not well established. Incidence of critical care therapy at time of index RC has been reported at 12.1%, though a majority of these patients underwent total parental nutrition initiation, which likely reflects sequelae of postoperative ileus and is typically temporary therapy.^29^ Association between dementia and intensive interventions after surgery remains mixed. Ottesen, *et al.* demonstrated that older adults with moderate-to-severe dementia undergoing elective total knee arthroplasty were at greater risk of requiring one or more intensive interventions; this effect was not observed among those with mild dementia.^30^ Conversely, Shah, *et al.* demonstrated that older adults with ADRD undergoing lower limb amputation for chronic limb-threatening ischemia were at significantly lower risk of undergoing intensive interventions.^31^ It is possible that our ADRD cohort reflects milder dementia and accordingly less strongly associated with severe post-RC complications that may require intensive interventions or reflects caregiver preferences to avoid additional interventions. While it has been demonstrated that feeding tube placement is not associated with improved survival among those with dementia,^6^ discussion around feeding tube placement after surgery will need to recognize the role for nutritional support in acute postoperative recovery.

We found that frail patients with and without concomitant ADRD tended to have comparably negative associations with burdensome TICs, SNF admission, and mortality, while robust patients with concomitant ADRD only exhibited a negative association with SNF admission. Shah, *et al.* by comparison found that every iteration of frail/robust and ADRD/non-ADRD patients were associated with poorer survival after lower limb amputation, though similarly found that frailty remained associated with postoperative care utilization regardless of ADRD status.^31^ The interaction between ADRD and frailty likely varies with disease process and unique surgical risks; that ADRD remains an independent risk for institutionalization after RC regardless of frailty highlights the need for preoperative geriatric assessment. Additionally, frailty and ADRD are both associated with poorer survival even after consideration of competing risks, which may reflect the impact of delaying care and more advanced disease at time of RC.^23^

Our study had several key limitations. We cannot describe dementia severity, which may impact outcomes; however, CFI has been shown to indicate moderate-to-severe ADRD for patients with dementia who are also frail.^32^ Additionally, we lack qualitative data on patient and caregiver decision-making as it pertains to transitions in care and pursuing intensive interventions. Lastly, we rely on claims-based proxies of dementia and frailty, which, while robustly validated, cannot replace a formal geriatric assessment.

## CONCLUSIONS

Among a contemporary, national cohort of older adults, preoperative frailty and dementia were associated with two or more TICs, SNF admission within a year of RC, and poorer survival. Frail patients were also more likely to undergo intensive interventions, including feeding tube placement and initiation of dialysis, within 90 days of cystectomy. Our work highlights the importance of preoperative geriatric assessments of cognitive and physiologic deficits and, when feasible, appropriate prehabilitation.

## Supporting information

Supplment: ICD Codes

## Data Availability

The data used in this study are available from the Centers for Medicare and Medicaid Services (CMS). Due to data use agreement restrictions, the analytic dataset cannot be shared directly. The analytic code is available from the corresponding author upon reasonable request.

## ABBREVIATIONS

ADRD: Alzheimer’s disease and related dementias
BC: Bladder cancer
CFI: Claims-based frailty index
CI: Confidence interval
ECI: Elixhauser Comorbidity Index
HCPCS: Healthcare Common Procedure Coding System
HR: Hazard ratio
IP: Inpatient
NAC: Neoadjuvant chemotherapy
NH: Nursing home
OR: Odds ratio
PLWD: Person living with dementia
RC: Radical cystectomy
SNF: Skilled nursing facility
TIC: Transition in care

## ACKNOWLEDGEMENTS

The authors have no acknowledgements.

## FUNDING

This research was supported by the National Institutes of Health’s National Institute on Aging, grant R01AG067507. The content is solely the responsibility of the authors and does not necessarily represent the official views of the National Institutes of Health’s National Institute on Aging.

## CONFLICTS OF INTEREST

The authors declare no competing financial interests or personal relationships that could have influenced the work reported in this paper.

## Notes

### Competing Interest Statement

The authors have declared no competing interest.

