## Supplementary Tables for "Dementia and Frailty Impact Postoperative Care Trajectories and Burden among Older Adults Undergoing Radical Cystectomy for Bladder Cancer"

Table 1a. Cumulative incidence of care trajectory outcomes based on univariable, unadjusted four-level variable analysis by ADRD/frailty status.

Table 1b. Cumulative incidence of care trajectory outcomes based on multivariable, adjusted four-level variable analysis by ADRD/frailty status.

Table 2a. Univariable unadjusted associations between ADRD status or frailty and outcomes.

Table 2b. Univariable unadjusted associations between four-level ADRD/frailty status and outcomes.

Table 3a. Univariable unadjusted competing risk and survival analysis between ADRD status or frailty and outcomes.

Table 3b. Univariable unadjusted competing risk and survival analysis between four-level ADRD/frailty status and outcomes.

Table 4a. Multivariable adjusted competing risk and survival analysis between ADRD status or frailty and outcomes.

Table 4b. Multivariable adjusted competing risk and survival analysis between four-level ADRD/frailty status and outcomes.

SUPPLEMENTARY TABLES

**Table 1a.** Cumulative incidence of care trajectory outcomes based on univariable, unadjusted four-level variable analysis by ADRD/frailty status.

|  | **ADRD/Frail** | | **ADRD/Robust** | | **non-ADRD/Frail** | | **non-ADRD/Robust** | |
| --- | --- | --- | --- | --- | --- | --- | --- | --- |
| **Outcome** | **%** | **95% CI** | **%** | **95% CI** | **%** | **95% CI** | **%** | **95% CI** |
| 90-day new mobility device (*N*=3600) | 14.5 | 8.3, 25.4 | 12.2 | 5.5, 26.8 | 14.2 | 11.0, 18.3 | 14.3 | 13.2, 15.6 |
| 1-year TIC (*N*=3524) | 49.7 | 40.9, 60.3 | 39.0 | 26.3, 58.0 | 54.0 | 47.9, 60.9 | 38.9 | 37.0, 41.0 |
| 30-day SNF admission, among community-dwelling patients (*N*=3429) | 41.8 | 33.8, 51.7 | 33.0 | 23.5, 46.4 | 34.6 | 30.9, 30.9 | 17.1 | 16.3, 17.9 |
| 90-day SNF admission (*N*=3429) | 46.6 | 38.1, 57.1 | 37.1 | 26.9, 51.2 | 38.9 | 34.1, 44.4 | 19.5 | 18.6, 20.5 |
| 180-day SNF admission (*N*=3429) | 48.7 | 41.5, 57.1 | 38.9 | 27.2, 55.6 | 40.7 | 35.4, 46.8 | 20.6 | 19.7, 21.5 |
| 365-day SNF admission (*N*=3429) | 51.8 | 43.8, 61.2 | 41.7 | 30.7, 56.5 | 43.6 | 38.1, 49.8 | 22.3 | 21.3, 23.3 |
| 30-day NH admission, among community-dwelling patients (*N*=3429) | 3.9 | 1.7, 8.6 | 3.5 | 1.2, 9.8 | 2.3 | 1.3, 3.9 | 1.2 | 0.9, 1.7 |
| 90-day NH admission (*N*=3429) | 6.4 | 3.0, 13.6 | 5.7 | 1.9, 16.8 | 3.8 | 2.3, 6.2 | 2.0 | 1.6, 2.5 |
| 180-day NH admission (*N*=3429) | 8.8 | 3.9, 20.1 | 7.9 | 3.0, 20.5 | 5.3 | 3.0, 9.2 | 2.8 | 2.3, 3.5 |
| 365-day NH admission (*N*=3429) | *No observed events at 365 days after discharge* | | | | | | | |
| 30-day SNF admission, among NH patients (*N*=98) | 38.3 | 24.3, 60.6 | 36.5 | 21.8, 61.1 | 50.9 | 43.4, 59.6 | 48.7 | 39.9, 59.5 |
| 90-day SNF admission (*N*=98) | 41.8 | 27.5, 63.5 | 39.8 | 24.6, 64.4 | 54.8 | 45.8, 65.6 | 52.6 | 43.7, 63.3 |
| 180-day SNF admission (*N*=98) | 43.0 | 27.6, 66.8 | 41.0 | 25.0, 67.2 | 56.1 | 47.0, 67.0 | 53.9 | 45.0, 64.6 |
| 365-day SNF admission (*N*=98) | *No observed events at 365 days after discharge* | | | | | | | |
| 90-day any intensive intervention (*N*=3600) | 6.2 | 3.1, 12.3 | 3.1 | 0.8, 11.6 | 7.5 | 5.6, 10.1 | 3.6 | 3.1, 4.3 |

ADRD = Alzheimer’s disease and related dementias; TIC = transitions in care; SNF = skilled nursing facility; NH = nursing home

**Table 1b.** Cumulative incidence of care trajectory outcomes based on multivariable, adjusted four-level variable analysis by ADRD/frailty status.

|  | **ADRD/Frail** | | **ADRD/Robust** | | **non-ADRD/Frail** | | **non-ADRD/Robust** | |
| --- | --- | --- | --- | --- | --- | --- | --- | --- |
| **Outcome** | **%** | **95% CI** | **%** | **95% CI** | **%** | **95% CI** | **%** | **95% CI** |
| 90-day new mobility device (*N*=3600) | 12.2 | 7.7, 19.2 | 10.2 | 5.1, 20.7 | 12.2 | 9.1, 16.3 | 12.9 | 11.3, 14.8 |
| 1-year TIC (*N*=3524) | 48.8 | 39.2, 60.8 | 40.2 | 26.1, 61.7 | 55.4 | 49.9, 61.5 | 41.8 | 38.5, 45.3 |
| 30-day SNF admission, among community-dwelling patients (*N*=3429) | 32.0 | 25.8, 39.6 | 27.1 | 19.0, 38.5 | 29.2 | 25.6, 33.3 | 16.6 | 14.9, 18.5 |
| 90-day SNF admission (*N*=3429) | 36.3 | 29.5, 44.7 | 30.8 | 21.7, 43.9 | 33.2 | 28.2, 39.2 | 19.1 | 17.2, 21.3 |
| 180-day SNF admission (*N*=3429) | 38.1 | 29.5, 49.3 | 32.5 | 22.7, 46.5 | 35.0 | 29.9, 40.9 | 20.2 | 18.3, 22.4 |
| 365-day SNF admission (*N*=3429) | 41.0 | 33.3, 50.4 | 35.1 | 26.8, 45.9 | 37.7 | 32.7, 43.4 | 22.0 | 20.2, 24.0 |
| 30-day NH admission, among community-dwelling patients (*N*=3429) | 3.2 | 1.4, 7.5 | 2.9 | 1.0, 8.5 | 2.1 | 1.2, 3.7 | 1.2 | 0.8, 1.7 |
| 90-day NH admission (*N*=3429) | 5.3 | 2.3, 12.0 | 4.9 | 1.7, 13.7 | 3.5 | 2.0, 6.0 | 2.0 | 1.4, 2.8 |
| 180-day NH admission (*N*=3429) | 7.4 | 3.2, 17.1 | 6.7 | 2.0, 22.7 | 4.9 | 0.0292, 0.0807 | 2.8 | 2.1, 3,6 |
| 365-day NH admission (*N*=3429) | *No observed events at 365 days after discharge* | | | | | | | |
| 30-day SNF admission, among NH patients (*N*=98) | 27.3 | 15.4, 48.2 | 24.1 | 12.2, 47.5 | 48.6 | 37.4, 63.1 | 43.9 | 33.9, 56.7 |
| 90-day SNF admission (*N*=98) | 30.2 | 16.6, 54.9 | 26.8 | 14.1, 51.0 | 52.9 | 40.6, 68.8 | 47.9 | 35.2, 65.1 |
| 180-day SNF admission (*N*=98) | 31.2 | 16.7, 58.3 | 27.7 | 12.1, 63.1 | 54.3 | 42.4, 69.5 | 49.3 | 36.7, 66.0 |
| 365-day SNF admission (*N*=98) | *No observed events at 365 days after discharge* | | | | | | | |
| 90-day any intensive intervention (*N*=3600) | 5.1 | 2.6, 10.1 | 2.9 | 0.8, 11.2 | 6.5 | 4.7, 9.0 | 3.8 | 3.0, 4.8 |

ADRD = Alzheimer’s disease and related dementias; TIC = transitions in care; SNF = skilled nursing facility; NH = nursing home

**Table 2a**. Univariable unadjusted associations between ADRD status or frailty and outcomes.

|  | ADRD | | | Frail | | |
| --- | --- | --- | --- | --- | --- | --- |
| Outcome | **OR** | **95% CI** | ***p*-value** | **OR** | **95% CI** | ***p*-value** |
| New mobility device (*N*=3309) | 1.27 | 0.75, 2.15 | 0.376 | 1.25 | 0.93, 1.69 | 0.139 |
| TIC (*N*=3220) | 1.97 | 1.25, 3.11 | 0.003 | 2.66 | 2.08, 3.41 | <.0001 |
| Functional Decline (*N*=3527) | 0.51 | 0.34, 0.76 | 0.001 | 0.34 | 0.28, 0.42 | <.0001 |
| 30-day SNF admission, among community-dwelling patients (*N*=3376) | 3.43 | 2.27, 5.20 | <.0001 | 3.22 | 2.51, 4.14 | <.0001 |
| 90-day SNF admission (*N*=3286) | 3.57 | 2.33, 5.47 | <.0001 | 3.90 | 3.03, 5.02 | <.0001 |
| 180-day SNF admission (*N*=3177) | 3.26 | 2.13, 4.99 | <.0001 | 3.85 | 2.99, 4.96 | <.0001 |
| 365-day SNF admission (*N*=2985) | 3.88 | 2.51, 5.99 | <.0001 | 3.88 | 2.98, 5.05 | <.0001 |
| 30-day NH admission, among community-dwelling patients (*N*=3347) | 4.72 | 1.47, 15.18 | 0.009 | 3.39 | 1.53, 7.51 | 0.003 |
| 90-day NH admission (*N*=3177) | 4.67 | 1.90, 11.51 | 0.001 | 3.28 | 1.72, 6.25 | 0.000 |
| 180-day NH admission (*N*=2996) | 3.27 | 1.35, 7.928 | 0.009 | 2.70 | 1.54, 4.72 | 0.001 |
| 365-day NH admission (*N*=2726) | 3.82 | 1.795, 8.134 | 0.001 | 2.67 | 1.62, 4.38 | 0.000 |
| 30-day SNF admission, among NH patients (*N*=91) | 0.81 | 0.245, 2.689 | 0.733 | 1.14 | 0.49, 2.64 | 0.764 |
| 90-day SNF admission (N=86) | 1.58 | 0.312, 8.058 | 0.579 | 1.43 | 0.57, 3.58 | 0.450 |
| 180-day SNF admission (*N*=83) | 1.30 | 0.253, 6.645 | 0.756 | 1.70 | 0.62, 4.66 | 0.300 |
| 365-day SNF admission (*N*=79) | 1.72 | 0.201, 14.746 | 0.621 | 3.12 | 0.94, 10.34 | 0.063 |
| 30-day mortality (*N*=3600) | 3.28 | 1.810, 5.940 | <.0001 | 3.44 | 2.33, 5.10 | <.0001 |
| 90-day mortality (*N*=3600) | 4.26 | 2.818, 6.450 | <.0001 | 3.73 | 2.91, 4.78 | <.0001 |
| 180-day mortality (*N*=3600) | 3.22 | 2.208, 4.685 | <.0001 | 3.36 | 2.73, 4.13 | <.0001 |
| 365-day mortality (*N*=3600) | 2.99 | 2.096, 4.258 | <.0001 | 2.78 | 2.30, 3.37 | <.0001 |
| 90-day intensive intervention (*N*=3314) | 2.50 | 1.269, 4.920 | 0.008 | 3.74 | 2.56, 5.48 | <.0001 |
| Any negative event (*N*=3600) | 1.56 | 0.904, 2.692 | 0.111 | 1.76 | 1.31, 2.37 | 0.000 |
| LOS* | 0.84 | 0.418, 2.106 | 0.190 | 3.23 | 2.25, 4.21 | <.0001 |

ADRD = Alzheimer’s disease and related dementias; TIC = transitions in care; SNF = skilled nursing facility; NH = nursing home; LOS = length of stay. Reference groups are non-ADRD and non-frail beneficiaries. *Outcomes for LOS reflects mean difference, not OR.

**Table 2b.** Univariable unadjusted associations between four-level ADRD/frailty status and outcomes.

|  | non-ADRD/Robust | ADRD/Frail | | | ADRD/Robust | | | non-ADRD/Frail | | |
| --- | --- | --- | --- | --- | --- | --- | --- | --- | --- | --- |
| Outcome |  | **OR** | **95% CI** | ***p*-value** | **OR** | **95% CI** | ***p*-value** | **OR** | **95% CI** | ***p*-value** |
| New mobility device (*N*=3309) | REF | 1.51 | 0.81, 2.82 | 0.194 | 1.08 | 0.55, 2.14 | 0.822 | 1.20 | 1.20, 1.65 | 0.258 |
| TIC (*N*=3220) | REF | 3.47 | 1.90, 6.32 | <.0001 | 0.78 | 0.36, 1.69 | 0.533 | 2.54 | 1.96, 3.28 | <.0001 |
| Functional Decline (*N*=3527) | REF | 0.38 | 0.23, 0.64 | 0.000 | 2.97 | 1.58, 5.60 | 0.001 | 0.33 | 0.27, 0.41 | <.0001 |
| 30-day SNF admission, among community-dwelling patients (*N*=3376) | REF | 4.61 | 2.71, 7.83 | <.0001 | 2.89 | 1.53, 5.46 | 0.001 | 3.04 | 2.32, 3.99 | <.0001 |
| 90-day SNF admission (*N*=3286) | REF | 5.21 | 3.04, 8.92 | <.0001 | 2.58 | 1.37, 4.85 | 0.003 | 3.74 | 2.84, 4.93 | <.0001 |
| 180-day SNF admission (*N*=3177) | REF | 4.85 | 2.81, 8.36 | <.0001 | 2.76 | 1.43, 5.31 | 0.002 | 3.74 | 2.84, 4.92 | <.0001 |
| 365-day SNF admission (*N*=2985) | REF | 6.20 | 3.50, 11.01 | <.0001 | 5.66 | 1.23, 26.01 | 0.026 | 3.59 | 2.70, 4.76 | <.0001 |
| 30-day NH admission, among community-dwelling patients (*N*=3347) | REF | 5.46 | 1.51, 19.70 | 0.010 | 4.77 | 1.34, 16.93 | 0.016 | 3.17 | 1.23, 8.18 | 0.017 |
| 90-day NH admission (*N*=3177) | REF | 5.73 | 2.02, 16.26 | 0.001 | 3.02 | 0.86, 10.54 | 0.083 | 2.95 | 1.44, 6.06 | 0.003 |
| 180-day NH admission (*N*=2996) | REF | 4.10 | 1.48, 11.39 | 0.007 | 3.14 | 1.03, 9.58 | 0.044 | 2.51 | 1.37, 4.61 | 0.003 |
| 365-day NH admission (*N*=2726) | REF | 5.12 | 2.10, 12.50 | 0.000 | 0.73 | 0.19, 2.76 | 0.641 | 2.33 | 1.35, 4.03 | 0.003 |
| 30-day SNF admission, among NH patients (*N*=91) | REF | 0.90 | 0.26, 3.11 | 0.863 | 1.35 | 0.24, 7.71 | 0.739 | 1.23 | 0.48, 3.15 | 0.665 |
| 90-day SNF admission (N=86) | REF | 1.81 | 0.34, 9.57 | 0.482 | 0.94 | 0.15, 5.78 | 0.949 | 1.35 | 0.50, 3.62 | 0.553 |
| 180-day SNF admission (*N*=83) | REF | 1.63 | 0.31, 8.6 | 0.568 | 0.78 | 0.07, 8.45 | 0.836 | 1.72 | 0.57, 5.23 | 0.337 |
| 365-day SNF admission (*N*=79) | REF | 2.57 | 0.29, 22.60 | 0.396 | 1.89 | 0.45, 8.0 | 0.386 | 3.30 | 0.88, 12.45 | 0.078 |
| 30-day mortality (*N*=3600) | REF | 4.99 | 2.61, 9.57 | <.0001 | 2.79 | 1.21, 6.42 | 0.016 | 3.13 | 1.99, 4.94 | <.0001 |
| 90-day mortality (*N*=3600) | REF | 6.45 | 4.02, 10.36 | <.0001 | 1.65 | 0.75, 3.62 | 0.211 | 3.26 | 2.47, 4.3 | <.0001 |
| 180-day mortality (*N*=3600) | REF | 5.17 | 3.35, 7.97 | <.0001 | 1.517 | 0.76, 3.02 | 0.235 | 3.03 | 2.38, 3.86 | <.0001 |
| 365-day mortality (*N*=3600) | REF | 4.75 | 3.13, 7.21 | <.0001 | 1.76 | 0.41, 7.47 | 0.445 | 2.45 | 1.98, 3.05 | <.0001 |
| 90-day intensive intervention (*N*=3314) | REF | 3.86 | 1.80, 8.26 | 0.001 | 0.12 | -2.09, 2.34 | 0.913 | 3.76 | 2.47, 5.73 | <.0001 |
| Any negative event (*N*=3600) | REF | 2.88 | 1.27, 6.55 | 0.012 | 0.63 | 0.32, 1.25 | 0.188 | 1.60 | 1.18, 2.17 | 0.003 |
| LOS* | REF | 1.72 | 0.22, 3.21 | 0.025 | 0.96 | 0.37, 2.48 | 0.926 | 3.60 | 2.41, 4.80 | <.0001 |

ADRD = Alzheimer’s disease and related dementias; TIC = transitions in care; SNF = skilled nursing facility; NH = nursing home; LOS = length of stay. Reference group is non-ADRD/robust beneficiaries. *Outcomes for LOS reflects mean difference, not OR.

**Table 3a**. Univariable unadjusted competing risk and survival analysis between ADRD status or frailty and outcomes.

|  | ADRD | | | Frail | | |
| --- | --- | --- | --- | --- | --- | --- |
| Outcome | **HR** | **95% CI** | ***p*-value** | **HR** | **95% CI** | ***p*-value** |
| New mobility device (*N*=3600) | 0.96 | 0.60, 1.52 | 0.851 | 1.00 | 0.76, 1.30 | 0.979 |
| TIC (*N*=3524) | 1.20 | 0.90, 1.59 | 0.215 | 1.54 | 1.31, 1.80 | <.0001 |
| SNF admission, among community-dwelling patients (*N*=3429) | 2.35 | 1.85, 2.99 | <.0001 | 2.34 | 1.98, 2.77 | <.0001 |
| NH admission, among community-dwelling patients (*N*=3429) | 2.89 | 1.42, 5.85 | 0.003 | 2.08 | 1.29, 3.36 | 0.003 |
| SNF admission, among NH patients (*N*=98) | 0.70 | 0.39, 1.28 | 0.249 | 0.97 | 0.68, 1.38 | 0.867 |
| Overall survival | 2.51 | 1.93, 3.28 | <.0001 | 2.43 | 2.09, 2.83 | <.0001 |
| Any intensive intervention (*N*=3600) | 1.31 | 0.70, 2.46 | 0.400 | 2.05 | 1.49, 2.82 | <.0001 |
| LOS (*N*=3600) | 0.80 | 0.67, 0.95 | 0.013 | 0.64 | 0.59, 0.70 | <.0001 |

ADRD = Alzheimer’s disease and related dementias; TIC = transitions in care; SNF = skilled nursing facility; NH = nursing home; LOS = length of stay. Reference groups are non-ADRD and non-frail beneficiaries.

**Table 3b.** Univariable unadjusted competing risk and survival analysis between four-level ADRD/frailty status and outcomes.

|  | non-ADRD/Robust | ADRD/Frail | | | ADRD/Robust | | | non-ADRD/Frail | | |
| --- | --- | --- | --- | --- | --- | --- | --- | --- | --- | --- |
| Outcome | **REF** | **HR** | **95% CI** | ***p*-value** | **HR** | **95% CI** | ***p*-value** | **HR** | **95% CI** | ***p*-value** |
| New mobility device (*N*=3600) | REF | 1.01 | 0.59, 1.75 | 0.962 | 0.84 | 0.36, 1.96 | 0.684 | 0.99 | 0.75, 1.31 | 0.943 |
| TIC (*N*=3524) | REF | 1.39 | 1.01, 1.92 | 0.045 | 1.00 | 0.59, 1.71 | 0.991 | 1.573 | 1.34, 1.85 | <.0001 |
| SNF admission, among community-dwelling patients (*N*=3429) | REF | 2.89 | 2.20, 3.79 | <.0001 | 2.14 | 1.41, 3.24 | 0.000 | 2.27 | 1.88, 2.73 | <.0001 |
| NH admission, among community-dwelling patients (*N*=3429) | REF | 3.24 | 1.44, 7.29 | 0.005 | 2.88 | 1.01, 8.27 | 0.049 | 1.89 | 1.10, 3.23 | 0.020 |
| SNF admission, among NH patients (*N*=98) | REF | 0.72 | 0.39, 1.34 | 0.304 | 0.68 | 0.36, 1.29 | 0.237 | 1.06 | 0.73, 1.55 | 0.743 |
| Overall survival | REF | 3.60 | 2.70, 4.81 | <.0001 | 1.45 | 0.81, 2.59 | 0.208 | 2.20 | 1.85, 2.62 | <.0001 |
| Any intensive intervention (*N*=3600) | REF | 1.75 | 0.87, 3.50 | 0.117 | 0.86 | 0.21, 3.52 | 0.832 | 2.12 | 1.50, 3.01 | <.0001 |
| LOS (*N*=3600) | REF | 0.64 | 0.52, 0.80 | <.0001 | 1.112 | 0.84, 1.47 | 0.451 | 0.64 | 0.59, 0.71 | <.0001 |

ADRD = Alzheimer’s disease and related dementias; TIC = transitions in care; SNF = skilled nursing facility; NH = nursing home; LOS = length of stay. Reference group is non-ADRD/robust beneficiaries.

**Table 4a.** Multivariable adjusted competing risk and survival analysis between ADRD status or frailty and outcomes.

|  | ADRD | | | Frail | | |
| --- | --- | --- | --- | --- | --- | --- |
| Outcome | **HR** | **95% CI** | ***p*-value** | **HR** | **95% CI** | ***p*-value** |
| New mobility device (*N*=3600) | 0.89 | 0.56, 1.43 | 0.639 | 0.94 | 0.71, 1.25 | 0.673 |
| TIC (*N*=3524) | 1.08 | 0.81, 1.44 | 0.602 | 1.44 | 1.23, 1.69 | <.0001 |
| SNF admission, among community-dwelling patients (*N*=3429) | 1.80 | 1.42, 2.30 | <.0001 | 1.92 | 1.60, 2.31 | <.0001 |
| NH admission, among community-dwelling patients (*N*=3429) | 2.42 | 1.16, 5.04 | 0.018 | 1.90 | 1.13, 3.22 | 0.016 |
| SNF admission, among NH patients (*N*=98) | 0.53 | 0.29, 0.95 | 0.034 | 0.93 | 0.65, 1.32 | 0.670 |
| Overall survival | 1.92 | 1.48, 2.50 | <.0001 | 1.81 | 1.53, 2.14 | <.0001 |
| Any intensive intervention (*N*=3600) | 1.07 | 0.56, 2.05 | 0.828 | 2.05 | 1.49, 2.82 | <.0001 |
| LOS (*N*=3600) | 0.84 | 0.70, 1.01 | 0.070 | 0.69 | 0.63, 0.76 | <.0001 |

ADRD = Alzheimer’s disease and related dementias; TIC = transitions in care; SNF = skilled nursing facility; NH = nursing home; LOS = length of stay. Reference groups are non-ADRD and non-frail beneficiaries.

**Table 4b.** Multivariable adjusted competing risk and survival analysis between four-level ADRD/frailty status and outcomes.

|  | non-ADRD/Robust | ADRD/Frail | | | ADRD/Robust | | | non-ADRD/Frail | | |
| --- | --- | --- | --- | --- | --- | --- | --- | --- | --- | --- |
| Outcome | **REF** | **HR** | **95% CI** | ***p*-value** | **HR** | **95% CI** | ***p*-value** | **HR** | **95% CI** | ***p*-value** |
| New mobility device (*N*=3600) | REF | 0.94 | 0.54, 1.64 | 0.826 | 0.78 | 0.33, 1.83 | 0.568 | 0.94 | 0.70, 1.26 | 0.668 |
| TIC (*N*=3524) | REF | 1.24 | 0.89, 1.73 | 0.207 | 0.95 | 0.56, 1.62 | 0.850 | 1.49 | 1.27, 1.76 | <.0001 |
| SNF admission, among community-dwelling patients (*N*=3429) | REF | 2.12 | 1.59, 2.84 | <.0001 | 1.74 | 1.18, 2.56 | 0.005 | 1.90 | 1.56, 2.32 | <.0001 |
| NH admission, among community-dwelling patients (*N*=3429) | REF | 2.71 | 1.15, 6.38 | 0.022 | 2.48 | 0.85, 7.20 | 0.096 | 1.76 | 0.99, 3.16 | 0.056 |
| SNF admission, among NH patients (*N*=98) | REF | 0.55 | 0.31, 1.00 | 0.049 | 0.48 | 0.24, 0.94 | 0.034 | 1.15 | 0.76, 1.75 | 0.499 |
| Overall survival | REF | 2.55 | 1.92, 3.39 | <.0001 | 1.23 | 0.69, 2.20 | 0.475 | 1.65 | 1.36, 2.00 | <.0001 |
| Any intensive intervention (*N*=3600) | REF | 1.37 | 0.66, 2.85 | 0.398 | 0.78 | 0.19, 3.19 | 0.728 | 1.74 | 1.20, 2.53 | 0.004 |
| LOS (*N*=3600) | REF | 0.68 | 0.54, 0.86 | 0.001 | 1.17 | 0.89, 1.55 | 0.265 | 0.69 | 0.63, 0.76 | <.0001 |

ADRD = Alzheimer’s disease and related dementias; TIC = transitions in care; SNF = skilled nursing facility; NH = nursing home; LOS = length of stay. Reference group is non-ADRD/robust beneficiaries.
